# Development of a Clinical Severity Score for Indian Sickle Cell Anaemia Patients

**DOI:** 10.1101/2025.10.07.25337518

**Authors:** Suraj S Nongmaithem, Amitabh Biswas, Swaroop Iyer, Jandhayala Vyshnavi, Archana Wath, Giriraj R Chandak, Dipty Jain

## Abstract

**Background:** Sickle cell anemia (SCA) is a common monogenic disorder but phenotypic heterogeneity is common among patients, especially in Indians. Scores to label them as per severity have mostly included non-Indians. We investigated the utility of existing pediatric severity score (PSS) in Indian patients and attempted to develop a severity score to facilitate informed management decisions.

**Materials and Methods:** A total of 171 SCA patients were recruited and two clinical experts categorized them into mild, moderate and severe groups based on clinical and biochemical parameters. We generated PSS and two other modified scores viz. Indian Severity Score 1 (ISS1) by including additional four clinical parameters, and Indian Severity Score 2 (ISS2) by replacing four biochemical measures with related clinical parameters. The patients were randomized and using the training set (N=86), severity scores cutoff values were decided and severity status was established in the testing set (N=85). Overall concordance between severity score-based and clinical expert categorization was calculated in three randomized sets.

**Results:** Using PSS, only 2/3^rd^ (66.7%) of Indian patients matched with the clinical assessment; the modified scores significantly improved the concordance; ISS1 (82.8%) and ISS2 (85.1%). Results were similar in all three random sets (80.0%-84.71% and 80.0%-88.2% for ISS1 and ISS2 respectively), suggesting robustness of modified scores in Indian patients. The highest concordance was observed in mild (81-100%) followed by severe (57-85%) groups in all three severity score models. The lowest concordance was observed in the moderate group (10-48%).

**Conclusions:** We have developed a robust population-specific score for identification of severity status in young Indian SCA patients. Inclusion of specific clinical symptoms in Indian patients underlines the importance of population-specific features to correctly categorize SCA patients. Further exploration of its utility in other populations is needed.

## Introduction

Sickle cell anemia (SCA) is caused by a single point mutation (p.Glu6Val) in the beta globin gene (*HBB*). The highest disease burden exists in India and the African countries including The Democratic Republic of the Congo, Nigeria (GBD 2021; Serjeant 2013). Although SCA is caused by a single mutation, the clinical manifestations and disease severity is highly heterogeneous, ranging from a near normal life to early mortality (Piel et al., 2017; Kato et al., 2018). Overall, disease complications can be categorized into two main groups: those primarily brought on by functional nitric oxide deficiency and hemolytic disease, which result in large vessel vasculopathy, leg ulcers, pulmonary hypertension, cerebrovascular disease, and nephropathy; and those due to vaso-occlusive ischemic events, which cause painful episodes and progressive organ damage, hyposplenism, osteonecrosis, and liver damage, etc., (Ware et al., 2017; Houwing et al., 2019). Considering the heterogeneity of disease manifestations, the management of SCA is a big challenge for the clinicians and difficult to cope for the patients. Thus, there is a need for better assessment of clinical parameters and development of a tool for prediction of future clinical course.

There are many important factors that help in understanding the disease pathogenesis and decide the course of disease (Platt et al., 1994; Bunn et al., 1997). Based on these factors, patients can be categorized as mild, moderate, and severe; thereby assisting the course of diagnosis and disease management (Ballas et al., 2010). Previous studies have attempted to design a method using available biochemical parameters and clinical events that can categorize SCA patients in different disease severity groups (Sebastiani et al., 2007; Tweel et al., 2010; Shah et al., 2020). Based on disease prognosis, Sebastiani et al., developed the disease severity score (DSS) model that can be used to compute a personalized disease severity score allowing therapeutic decisions to be made (Sebastiani et al., 2007). A pediatric score system (PSS) developed using 12 parameters (8 clinical and 4 biochemical parameters) has shown significant differences in the scores for mild, moderate, and severely affected SCA patients (Table 1) (Tweel et al., 2010). However, these scores have not been evaluated in Indian SCA patients and no such score exists for them.

**Table 1.** Parameters included in different severity score for Sickle Cell Anaemia patients and their respective weights.

| Parameter groups | Parameters | PSS | ISS1 | ISS2 | Score weight |  |  |
| --- | --- | --- | --- | --- | --- | --- | --- |
|  |  |  |  |  | Absent | Present |  |
| Lifetime cumulative incidence | Bone Necrosis (avascular) | yes | yes | yes | 0 | 10 |  |
|  | Cerebral infarcts or vasculopathy | yes | yes | yes | 0 | 50 |  |
|  | Hepatic sequestration (acute) | yes | yes | yes | 0 | 50 |  |
|  | Pneumococcal meningitis/septicemia | yes | yes | yes | 0 | 50 |  |
|  | Priapism | yes | yes | yes | 0 | 10 |  |
|  | Splenic sequestration (acute) | yes | yes | yes | 0 | 50 |  |
| Laboratory values | Haemoglobin (0 if > 6.6 g dl<sup>-1</sup>) | yes | yes | no | 0 | 5 |  |
|  | Fetal Hemoglobin (0 if > 3.0%) | yes | yes | no | 0 | 5 |  |
|  | Lactate dehydrogenase (0 if ≤ 700 U/L) | yes | yes | no | 0 | 5 |  |
|  | Leucocytes (0 if < 15.2x10<sup>9</sup>/L) | yes | yes | no | 0 | 5 |  |
|  |  |  |  |  |  | 1- 2 episodes | >2 episodes |
| Number of recurrences over past 2 years | Acute chest syndrome | yes | yes | yes | 0 | 10 | 20 |
|  | Painful crisis | yes | yes | yes | 0 | 5 | 10 |
| New parameters (Number of recurrences over past 2 years) | Blood transfusion | no | yes | yes | 0 | 5 | 10 |
|  | Severe anaemia | no | yes | yes | 0 | 5 | 10 |
|  | Acute febrile illness | no | yes | yes | 0 | 5 | 10 |
|  | Others<sup>#</sup> | no | yes | yes | 0 | 5 | 10 |

PSS, pediatric severity score developed by Tweel et al. ISS1, Indian severity score model 1; ISS2, Indian severity score model 2. <sup>#</sup> includes cardiac myopathy, pulmonary hypertension, intestinal obstruction, severe infections, hemolytic crisis, acute hepatic failure, hypersplenism, renal failure, meningoencephalitis, encephalopathy, seizures, leg ulcers, hand foot syndrome, cholelithiasis, acute gastroenteritis, pneumonia, respiratory tract infections.

This study was initiated with an aim to develop a method that may predict the severity outcome of Indian SCA patients to assist the clinician in better disease management. We applied the PSS on young Indian SCA patients and also modified various clinical and biochemical parameters included in the score, as suggested by expert clinicians based on their long-term experience with the clinical phenotype, disease course and management of SCA patients. The modified scores were significantly better in identifying the severity status of the patients confirming the need for a population-specific severity score for better care of SCA patients.

## Methods

### Patient Recruitment and Data Collection

SCA patients attending the Outpatient Department of Government Medical College and Hospital, Nagpur were recruited after obtaining the written informed consent. Informed consent was taken from all the patients and the study was approved by the Institutional Ethics Committee of CSIR-CCMB (No.IEC-65-R1/2019, dated 10.03.2021). The inclusion criteria were patients aged 0 to18 years, diagnosed with SCA based on the SS pattern on high performance liquid chromatography analysis. A total of 171 patients (82 females and 89 males) were evaluated for the severity index. No biological sample was freshly collected for any investigations related to this study. Medical records from the hospital were utilized to collate the clinical data and laboratory investigations. Details of clinical parameters used for generation of the severity score are given in Table 1 and clinical characteristics of the patients are shown in Table 2. The number of recurrences of SCA-related complications was documented over last two years. Based on their prior experience and data from the patient medical records, two seasoned pediatricians (DJ and SI) with expertise in SCA independently classified the patients into three groups; mild, moderate, and severe. Any differences of opinion between the two experts was discussed and final decision was made with mutual agreement.

**Table 2:**
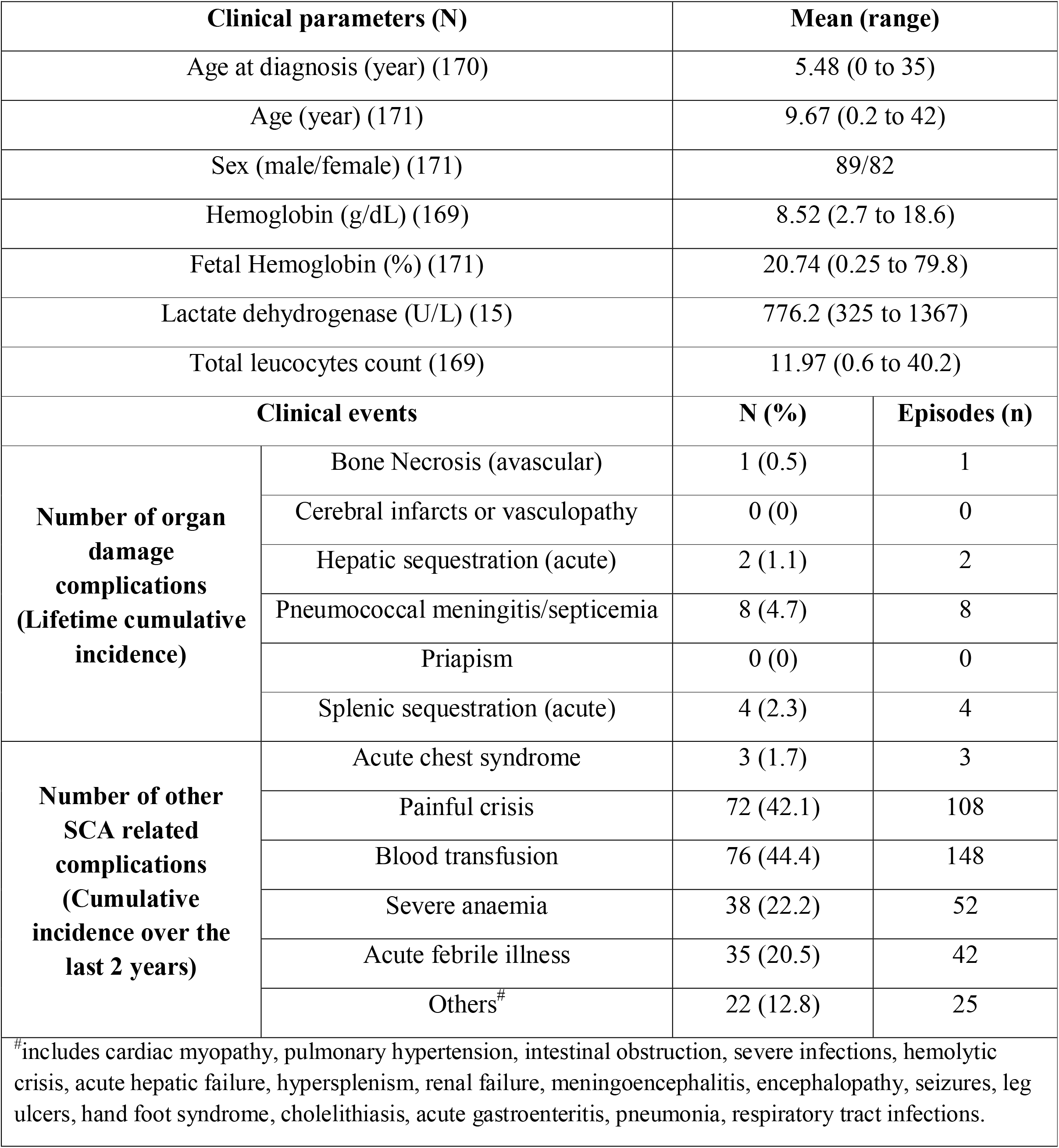
Clinical characteristic of Sickle Cell Anemia patients used in the study.

### Evaluation and validation of Severity Score

We first generated the PSS using best-weighted system including 12 clinical parameters (Table 1) in all 171 patients and tested its ability to categorize patients with respect to the clinical experts’ classification. Out of the 12 clinical parameters, 6 were related to cumulative lifetime incidence of organ damage, 2 other parameters were related to the number of recurrences of SCA-related complications over 2 years and, 4 were laboratory test values (Table 1). Out of three weight scoring system – Scores A, B and C (Tweel et al. 2010), we adopted Score C weight system where weights (range = 5 to 50 points) were assigned to the clinical parameters, according to the frequency and severity of clinical complications.

Based on the generated score, patients were categorized into mild, moderate, and severe and compared with clinical categorization as mentioned above.

We modified the PSS by incorporating 4 additional clinical parameters to the existing 12 features and termed it as Indian Severity Score 1 (ISS1) (Table 1). Weights were ascribed to each additional parameter based on the number of recurrences over last 2 years. For instance, patients with no history of blood transfusion in last 2 years were assigned the weight of ‘0’, while the weight was ‘5’ for patients with 1 to 2 episodes, and ‘10’ for more than 2 episodes of blood transfusions. Likewise, other parameters were also weighted as per the patients’ clinical profile. Considering the fact that the biochemical parameters in SCA are sensitive to clinical status of the patients and/or environmental factors, in the next step, we replaced all the biochemical parameters with relevant clinical parameters (8 from PSS and 4 additional parameters as mentioned in Table 1) and generated the Indian Severity Score 2 (ISS2).

We randomized the 171 SCA patients using *rand()* formula in the Excel and divided them into two groups, training group (N=86) and testing group (N=85). Further, the patients in both groups were randomized into three independent sets – R1, R2 and R3. We calculated descriptive statistics of three scoring systems-PSS, ISS1 and ISS2 in clinically classified mild, moderate and severe category of patients in the training group. Based on the median and inter-quartile ranges (IQR) from three random training sets, we defined the cutoff score values of PSS, ISS1 and ISS2 for categorization into mild, moderate and severe groups (Table 3). The cutoff score values of respective score systems were applied to the testing group (N=85) and the patients were categorized as per the severity status. The severity score-based groups were compared with clinical expert categorized respective groups independently in each random testing set (Table 4). The overall concordance of severity score-based and clinical expert categorization was calculated by combining the results from all three random tests for each score.

**Table 3:** Definition of cutoffs from descriptive statistics of different severity scores in Indian SCA patients (N=86; three random training sets)

| Model | Mild |  |  | Moderate |  |  | Severe |  |  |
| --- | --- | --- | --- | --- | --- | --- | --- | --- | --- |
|  | N | Mean (SD) | Median (IQR) | N | Mean (SD) | Median (IQR) | N | Mean (SD) | Median (IQR) |
| PSS_R1 | 52 | 3.27 (4.30) | 0 (0-5) | 15 | 5.33 (4.42) | 5 (0-10) | 19 | 30 (24.49) | 20 (10-55) |
| PSS_R2 | 54 | 2.96 (3.57) | 0 (0-5) | 13 | 5.77 (4.49) | 5 (5-10) | 19 | 22.11 (21.88) | 15 (5-50) |
| PSS_R3 | 62 | 3.47 (4.30) | 0 (0-5) | 10 | 6.5 (4.74) | 5 (5-10) | 14 | 28.93 (26.62) | 20 (5-55) |
| **PSS cutoff** | **0-5** |  |  | **>5-10** |  |  | **>10** |  |  |
| ISS1_R1 | 52 | 6.44 (5.08) | 5 (5-10) | 15 | 14.33 (3.20) | 15 (10-15) | 19 | 39.74 (23.72) | 35 (20-65) |
| ISS1_R2 | 54 | 5.19 (5.04) | 5 (0-10) | 13 | 15 (4.08) | 15 (15-15) | 19 | 32.89 (20.57) | 25 (20-55) |
| ISS1_R3 | 62 | 6.37 (5.29) | 5 (0-10) | 10 | 15.5 (4.38) | 15 (15-15) | 14 | 38.93 (24.35) | 32.5 (15-60) |
| **ISS1 cutoff** | **0-10** |  |  | **>10-15** |  |  | **>15** |  |  |
| ISS2_R1 | 52 | 4.81 (3.70) | 5 (0-7.5) | 15 | 12.67 (3.20) | 10 (10-15) | 19 | 35.79 (22.87) | 30 (15-55) |
| ISS2_R2 | 54 | 4.17 (3.85) | 5 (0-5) | 13 | 12.69 (3.30) | 10 (10-15) | 19 | 28.42 (19.79) | 15 (15-55) |
| ISS2_R3 | 62 | 4.76 (3.89) | 5 (0-10) | 10 | 12 (2.58) | 10 (10-15) | 14 | 35 (21.93) | 27.5 (15-55) |
| **ISS2 cutoff** | **0-10** |  |  | **>10-15** |  |  | **>15** |  |  |

SCA, sickle cell anaemia; PSS- pediatric severity score; ISS1- Indian severity score 1; ISS2- Indian severity score 2; R1, R2 and R3 are random training sets 1, 2 and 3 respectively; SD- standard deviation; IQR-inter quartile range

**Table 4:** Concordance between different severity scores and the clinically categorized groups of Indian SCA patients (N=85; from three random testing sets)

| Pediatric severity index (PSS) |  |  |  |  |  |  |  |  |  |  |
| --- | --- | --- | --- | --- | --- | --- | --- | --- | --- | --- |
| Clinical | Random Set 1 |  |  | Random Set 2 |  |  | Random Set 3 |  |  | Concordance (%) |
|  | Mild | Moderate | Severe | Mild | Moderate | Severe | Mild | Moderate | Severe |  |
| Mild (N=57) | 47 | 9 | 1 | 43 | 9 | 3 | 40 | 6 | 1 | 130/159 (81.76%) |
| Moderate (N=8) | 6 | 1 | 1 | 8 | 1 | 1 | 11 | 1 | 1 | 3/31 (9.68%) |
| Severe (N=20) | 6 | 3 | 11 | 5 | 3 | 12 | 5 | 6 | 14 | 37/65 (56.92) |
| Overall (%) | 59/85 (69.41) |  |  | 56/85 (65.88) |  |  | 55/85 (64.71) |  |  | 170/255 (66.67%) |
| Indian severity score 1 (ISS1) |  |  |  |  |  |  |  |  |  |  |
| Clinical | Random Set 1 |  |  | Random Set 2 |  |  | Random Set 3 |  |  | Concordance (%) |
|  | Mild | Moderate | Severe | Mild | Moderate | Severe | Mild | Moderate | Severe |  |
| Mild (N=57) | 52 | 3 | 2 | 48 | 5 | 2 | 43 | 3 | 1 | 143/159 (89.94) |
| Moderate (N=8) | 1 | 3 | 4 | 2 | 4 | 4 | 3 | 6 | 4 | 13/31 (41.94) |
| Severe (N=20) | 1 | 2 | 17 | 3 | 1 | 16 | 2 | 1 | 22 | 55/65 (84.62) |
| Overall (%) | 72/85 (84.71) |  |  | 68/85 (80.00) |  |  | 71/85 (83.53) |  |  | 211/255 (82.75) |
| Indian severity score 2 (ISS2) |  |  |  |  |  |  |  |  |  |  |
| Clinical | Random Set 1 |  |  | Random Set 2 |  |  | Random Set 3 |  |  | Concordance (%) |
|  | Mild | Moderate | Severe | Mild | Moderate | Severe | Mild | Moderate | Severe |  |
| Mild (N=57) | 57 | 0 | 0 | 55 | 0 | 0 | 47 | 0 | 0 | 159/159 (100.00) |
| Moderate (N=8) | 3 | 4 | 1 | 4 | 5 | 1 | 5 | 6 | 2 | 15/31 (48.39) |
| Severe (N=20) | 1 | 6 | 13 | 3 | 2 | 15 | 2 | 8 | 15 | 43/65 (66.15) |
| Overall (%) | 74/85 (87.10) |  |  | 75/85 (88.24) |  |  | 68/85 (80.00) |  |  | 217/255 (85.10) |

SCA, sickle cell anaemia; PSS- pediatric severity score; ISS1- Indian severity score 1; ISS2- Indian severity score 2; R1, R2 and R3 are random training sets 1, 2 and 3 respectively.

## Results

### Patient characteristics

The clinical characteristics of the patients are given in Table 2. The average age at recruitment was 9.7 years. Instances of vaso-occlusive or painful crises (VOC) and recurrent blood transfusion were two most common complaints in majority of the patients (Table 2). VOC was reported in 72 SCA patients and blood transfusion in 76 SCA patients. Repeated hospitalizations due to severe anemia (N=38) and acute febrile illness (N=35) were also very high among SCA patients (Table 2).

Out of 171 patients, the clinical experts classified 109 (63.7%) patients as belonging to mild, 23 (13.5%) to moderate, and 39 (22.8%) to severe category. In three randomized training sets comprising of 86 SCA patients each, proportion of mild (60.5%, 62.8% and 72.1%), moderate (17.4%, 15.1% and 11.6%) and severe (22.1%, 22.1% and 16.3%) categories were overlapping (Table 3). Based on the median (IQR), we defined the PSS cutoff value of 0 to 5 as mild, >5 to 10 as moderate, and >10 as severe category (Table 3), which were applied to the testing set of 85 patients. Distribution of SCA patients in three severity categories was checked for concordance with clinical expert classification. Overall from three random sets of SCA patients, a 66.67% concordance was noted between PSS-based categorization and the clinical expert classification (Table 4). The highest concordance was observed in mild (81.8%), followed by severe (56.9%), and lowest in the moderate category (9.68%).

Based on the clinical relevance in Indian SCA patients, 4 additional clinical features were added to 12 parameters included in the PSS and a new score ISS1 was generated (Table 1 and Methods). For ISS1, a cutoff value of 0 to 10 as mild, >10 to 15 as moderate, and >15 as severe (Table 3) was specified and applied to the testing set patients. The overall concordance between ISS1-based and clinical expert defined categorization significantly improved to 82.75% (Table 4). While concordance in the mild category improved marginally (89.9%), a significant improvement was noted both in severe (84.6%) and the moderate group (41.9%) (Table 4).

Since the biochemical parameters are sensitive to clinical state of the patient (steady vs crisis state) and other environmental factors, we replaced the four biochemical parameters in ISS1 with related clinical features and generated a new score ISS2 (Table 1). Using same cut off value points as for ISS1, we observed a marginal improvement for ISS2 in overall concordance with clinical expert classification (85.1%) (Table 3). Interestingly, a significant improvement was noted for mild (100%) and moderate (48.4%) categories, a substantial drop of ∼18% was noted for the severe group of patients (66.1%) (Table 4).

## Discussion

Sickle cell anaemia is a global genetic disorder due to a single mutation in the beta globin gene; yet phenotypic heterogeneity is very common, making patient management challenging. Multiple severity scores have been proposed for SCA, combining clinical features and laboratory parameters to make a meaningful single entity that can confidently measure morbidity and/or mortality within a given time. Among several such, two scores, namely Disease Severity Score (DSS) (Sebastiani et al., 2007) and Pediatric Severity Score (PSS) (Tweel et al., 2010) are commonly used, of which, PSS is considered the most appropriate for SCD children of other ancestries. Differences in the clinical presentation and disease severity in patients from different ancestral populations demands population-specific severity scores (Steinberg et al., 2012; Saraf et al., 2014; Seth et al., 2025). We tested the PSS in young Indian SCA patients and observed that it functions poorly in proper annotation of their severity status. Modified scores based on inclusion of additional clinical features prevalent in the Indian patients significantly improved the predictability. Further replacement of various hematological parameters with related clinical features made it more robust. Overall, we demonstrate that severity scores based on population-specific clinical features allow more accurate identification of patients’ severity status and thus empower the clinicians to design the disease management.

One of the first attempts to identify manifestations that can predict severe clinical course in SCA infants was made by Miller et al, who identified dactylitis, severe anaemia and leukocytosis as three easily identifiable features (Miller et al, NEJM 2000). However, it was Sebastini, who developed the first severity score predicting the risk of death within 5 years (Sebastini et al., 2007). Pediatric severity score (PSS) developed by Tweel et al., discriminates young SCA patients into mild, moderate and severe categories that could help tailor the management and hydroxyurea therapy (Tweel et al, 2010). We observed that the PSS could correctly determine the severity status in only 2/3^rd^ of Indian SCA children, suggesting that certain unique clinical/biochemical parameters in Indian patients might be restricting the applicability of PSS for determining severity in Indian SCA patients. This could be related to very low occurrence of organ damage complications which are assigned high scores in the PSS system, as identified in this study. Analyzing the frequency of clinical features and lifetime cumulative incidence of long-term complications, we observed the former to be highly variable in SCA patients in other ancestries. While recurrent painful crises was the most common symptom as in other populations, recurrent blood transfusion was another common consequence (Seth et al., 2025). Further, severe anemia and acute febrile sickness led to repeated hospitalizations in Indian SCA patients (Table 2). The inclusion of four frequently observed clinical features in Indian SCA patients to the PSS (referred as Indian severity score 1) significantly improved the discrimination as per severity, confirming that these clinical features hold larger importance in Indian patients. We further argued that an only clinical features-based score would empower the clinician to take immediate management decision, since loss to follow-up is a common problem. Hence, we replaced the four biochemical parameters – total hemoglobin, fetal hemoglobin, lactate dehydrogenase and total leucocyte count included in the original PSS with clinical features representing the same status. The All Clinical parameters-based severity score, Indian Severity Score 2 performed marginally better than the combined clinical and laboratory parameters-based Indian Severity Score 1 and was highly concordant with the clinical expert classification. This indicates that population-specific clinical parameters which are routinely recorded during clinical examination can be used to generate severity scores which can predict the future risk of clinical complication and mortality risk.

While both ISS1 and ISS2 provide an overall similar concordance with the clinical experts’ categorization, it is interesting to note that the ISS2 correctly predicts all the mild cases while ISS1 performs much better in predicting severe cases. The concordance in severe cases drops by close to 20%, which has important implications as early intervention with hydroxyurea and regular follow up is the key to ensuring a good quality of life for SCA patients. This also confirms the established convention that biochemical and hematological parameters are better indicators of disease severity in SCA patients. However, it may be argued that all the misclassifications are between moderate and severe cases; both categories have largely similar management plan and hence the difference between ISS1 and ISS2-based grouping of the patients may not significantly impact the management decision. Overall, the ISS1 can help the clinicians to decide the management on their first contact and further qualified by inclusion of the hematological status of the patients, which are anyways required for monitoring the hydroxyurea therapy.

The study has several strengths and few limitations. This is the first study on generation of clinical severity score based on long-term observations on clinical symptoms and course in Indian SCA patients. Developing from an established score and involvement of SCA clinical experts, personally involved in the follow-up of each patient brings quality to these assessments. One of the limitations of the study is that a significant proportion of data is collected from the records, however, involvement of well-trained pediatricians ensures data integrity and robustness. Another limitation was missing LDH levels in majority of the patients, although all other parameters used in the pediatric severity score were available with us. It may also be noted that the severity scores have been generated on patients from one region of India and clinical differences in patients from other regions in the country cannot be ruled out (Hockham et al., 2018).

To summarize, we have developed a population-specific severity score suited for sickle cell anemia patients in Indians which can be used by clinicians to predict disease severity. However, systematic studies to evaluate its performance in independent cohorts from different regions of the country is needed to validate its wider utilization in predicting the course of the disease. This can help the clinicians in better disease management and modulate future clinical course in the patients.

## Data Availability

All data produced in the present study are available upon reasonable request to the authors

## Acknowledgements

The authors acknowledge all sickle cell anaemia patients and their families who, despite their sufferings participated in the study. Thanks are also due to the staff of Department of Pediatrics, Government Medical College and Hospital and Arihant Superspeciality Hospital, Nagpur for their continuous support in managing the patients. Special mention is made for the CSIR-CCMB staff who traveled to long distance and contributed in various patient care camps during the course of the CSIR-Sickle Cell Anaemia project.

## References

1. Ballas SK, Lieff S, Benjamin LJ, Dampier CD, Heeney MM, Hoppe C, Johnson CS, Rogers ZR, Smith-Whitley K, Wang WC, Telen MJ; Investigators, Comprehensive Sickle Cell Centers. Definitions of the phenotypic manifestations of sickle cell disease. Am J Hematol. 2010 Jan;85(1):6–13. doi: 10.1002/ajh.21550.

2. Bunn HF. Pathogenesis and treatment of sickle cell disease. N Engl J Med. 1997 Sep 11;337(11):762–9. doi: 10.1056/NEJM199709113371107.

3. GBD 2021 Sickle Cell Disease Collaborators. Global, regional, and national prevalence and mortality burden of sickle cell disease, 2000-2021: a systematic analysis from the Global Burden of Disease Study 2021. Lancet Haematol. 2023 Aug;10(8):e585–e599. doi: 10.1016/S2352-3026(23)00118-7.

4. Hockham C, Bhatt S, Colah R, Mukherjee MB, Penman BS, Gupta S, Piel FB. The spatial epidemiology of sickle-cell anaemia in India. Sci Rep. 2018 Dec 6;8(1):17685. doi: 10.1038/s41598-018-36077-w.

5. Houwing ME, de Pagter PJ, van Beers EJ, Biemond BJ, Rettenbacher E, Rijneveld AW, Schols EM, Philipsen JNJ, Tamminga RYJ, van Draat KF, Nur E, Cnossen MH; SCORE Consortium. Sickle cell disease: Clinical presentation and management of a global health challenge. Blood Rev. 2019 Sep;37:100580. doi: 10.1016/j.blre.2019.05.004.

6. Kato GJ, Piel FB, Reid CD, Gaston MH, Ohene-Frempong K, Krishnamurti L, Smith WR, Panepinto JA, Weatherall DJ, Costa FF, Vichinsky EP. Sickle cell disease. Nat Rev Dis Primers. 2018 Mar 15;4:18010. doi: 10.1038/nrdp.2018.10.

7. Miller ST, Sleeper LA, Pegelow CH, Enos LE, Wang WC, Weiner SJ, Wethers DL, Smith J, Kinney TR. Prediction of adverse outcomes in children with sickle cell disease. N Engl J Med. 2000 Jan 13;342(2):83–9. doi: 10.1056/NEJM200001133420203.

8. Piel FB, Steinberg MH, Rees DC. Sickle Cell Disease. N Engl J Med. 2017 Apr 20;376(16):1561–1573. doi: 10.1056/NEJMra1510865.

9. Platt OS, Brambilla DJ, Rosse WF, Milner PF, Castro O, Steinberg MH, Klug PP. Mortality in sickle cell disease. Life expectancy and risk factors for early death. N Engl J Med. 1994 Jun 9;330(23):1639–44. doi: 10.1056/NEJM199406093302303.

10. Saraf SL, Molokie RE, Nouraie M, Sable CA, Luchtman-Jones L, Ensing GJ, Campbell AD, Rana SR, Niu XM, Machado RF, Gladwin MT, Gordeuk VR. Differences in the clinical and genotypic presentation of sickle cell disease around the world. Paediatr Respir Rev. 2014 Mar;15(1):4–12. doi: 10.1016/j.prrv.2013.11.003.

11. Sebastiani P, Nolan VG, Baldwin CT, Abad-Grau MM, Wang L, Adewoye AH, McMahon LC, Farrer LA, Taylor JG 6th, Kato GJ, Gladwin MT, Steinberg MH. A network model to predict the risk of death in sickle cell disease. Blood. 2007 Oct 1;110(7):2727–35. doi: 10.1182/blood-2007-04-084921.

12. Serjeant GR. The natural history of sickle cell disease. Cold Spring Harb Perspect Med. 2013 Oct 1;3(10):a011783. doi: 10.1101/cshperspect.a011783.

13. Seth T, Udupi S, Jain S, Bhatwadekar S, Menon N, Jena RK, Kumar R, Ray S, Parmar B, Goel AK, Vasava A, Dutta A, Samal P, Ballikar R, Bhat D, Dolai TK, Bhattacharyya J, Shetty D, Mistry M, Jain D. Burden of vaso-occlusive crisis, its management and impact on quality of life of Indian sickle cell disease patients. Br J Haematol. 2025 Jan;206(1):296–309. doi: 10.1111/bjh.19829.

14. Shah N, Beenhouwer D, Broder MS, Bronte-Hall L, De Castro LM, Gibbs SN, Gordeuk VR, Kanter J, Klings ES, Lipato T, Manwani D, Scullin B, Yermilov I, Smith WR. Development of a Severity Classification System for Sickle Cell Disease. Clinicoecon Outcomes Res. 2020 Oct 28;12:625–633. doi: 10.2147/CEOR.S276121.

15. Steinberg MH, Sebastiani P. Genetic modifiers of sickle cell disease. Am J Hematol. 2012 Aug;87(8):795–803. doi: 10.1002/ajh.23232.

16. van den Tweel XW, van der Lee JH, Heijboer H, Peters M, Fijnvandraat K. Development and validation of a pediatric severity index for sickle cell patients. Am J Hematol. 2010 Oct;85(10):746–51. doi: 10.1002/ajh.21846.

17. Ware RE, de Montalembert M, Tshilolo L, Abboud MR. Sickle cell disease. Lancet. 2017 Jul 15;390(10091):311–323. doi: 10.1016/S0140-6736(17)30193-9.

